# Deep Learning in Opthalmology: Iris Melanocytic Tumors Intelligent Diagnosis

**DOI:** 10.1101/2021.09.14.21263573

**Authors:** Abdulkader Helwan

## Abstract

Recently, Convolutional neural networks (CNN) have shown a growth due to their ability of learning different level image representations that helps in image classification in different fields. These networks have been trained on millions of images, so they gained a powerful ability of extracting the rightful features from input images, which results in accurate classification. In this research, we investigate the effects of transfer learning based convolutional neural networks for the iris tumor malignancy identification as it is notoriously hard to distinguish an iris nevus from an iris tumor. Features are transferred from a CNN trained on a source task, i.e. ImageNet, to a target task, i.e. iris tumor datasets. We transfer features learned from AlexNet and VGG-16 that are trained on ImageNet, to classify three different iris images types which are: iris nevus unaffected, iris cysts, and iris melanocytic tumors. The employed pre-trained models are modified by replacing their feedforward neural network classifier, Softmax, by a support vector machine (SVM) that is expected to slightly boost their performance (AlexNet-SVM and VGG16-SVM). All employed models are trained (fine-tuned) on a 60% of the available large dataset of iris images in order to investigate their power of generalization when trained using large amount of data. The networks are also tested on 40% of the data. The best performance was achieved by the VGG16-SVM which scored a high accuracy of 96.27% and strong features extraction capability as compared to the other models. Experimentally, it was seen that adding SVM contributed in improving the network performance compared to original models which use a feedforward neural network classifier.

## Introduction

Iris tumors have a wide spectrum, starting from nevus to melanoma to metastasis. Few comprehensive studies explained this array of iris tumors, which were mostly laboratory pathology studies [1,2]. Generally, iris tumors are broadly classified into two main categories: cystic lesions and melanocytic tumors of the iris. The cystic lesions or the iris include two main cysts which are stromal cysts and iris pigment epithelial (IPE) cysts. Stromal cysts are characterized by a smooth surface and a transparent mass on or within the iris stroma. Note that stromal cysts can be either congenital or acquired. This tumor can lead to more serious effects such as secondary iritis, photophobia, pain and glaucoma.

The IPE cysts are the lesions arisen from the surface of the iris and they used to be misdiagnosed with the ciliary body melanoma due to their similarities in terms of color and pattern. These IPE cysts can be sorted into mid-zonal (21%), margin (3%), also peripheral (73%), free floating (<1%), and dislodged (3%) [1].

The second category of iris tumors is the iris melanocytic tumors which are benign tumors with tendency to malignancy transformation. Iris melanocytic tumors consists of the nevus, freckle, melanocytoma, and melanoma. Note that these tumors are benign except the melanoma, which is considered a malignant tumor. According to many medical reports [3,4] which stated that benign iris tumors sometimes transform into iris melanoma, which is malignant iris tumor and hence requires an urgent medical interference.

The identification of the iris tumor type is still a wearying process for ophthalmologists. Thus, the advances in computer systems that aids in diagnosing the iris melanocytic tumors is an advancement of dire need. Recently, deep neural networks have been significantly useful in solving different types of medical conditions [5,6]. Deep learning is a process in machine learning models that consists of a deep structure granting it a great ability in attaining the mid and high levels of abstractions from input raw data [7].

Recently, deep neural networks, in specific convolutional neural networks (CNNs), have gained substantial attention by scientists in the medical field, owing to its great efficiency in image classification [5,8]. This inspired scientists to transfer the knowledge attained by these deep networks when trained on millions of images to address medical image diagnosis and classification tasks. Deep learning models are mostly effective when a large training dataset is applied. In the medical field, large datasets are not usually available. Hence, transfer learning may be the only solution. Transfer learning refers to the use of pre-trained convolutional neural networks that are already trained on large datasets such as ImageNet [9], and benefit from their learned parameters, in specific weights, to the destination target network model.

Those deep networks can lead to the same obstacles and drawbacks as a feedforward neural network can do. This is due to the same training algorithm they both use, which is the conventional back propagation algorithm.

For a neural network to be effective, it should perform significantly efficient during training and testing the datasets. This means that the variance error and bias error should maintain a good balance [10]. Otherwise, a problem called underfitting will reveal if a high bias and low variance situation occur. It should be noted also that some more complex models may encounter another problem called overfitting if a high variance and low bias occur. These two drawbacks are considered major problems when training a complex feedforward neural network.

In order to alleviate these problems, several approaches were proposed [11]. Some of these approaches propose that weights pre-training, early training stopping, and dropping out some hidden neurons can allow a network to reach a good fit.

However, in this study we explore whether or not these problems can be avoided by switching the Softmax in neural network with a multi-class SVM that is supposed to act as a classifier for both the pre-trained models used. Note that this solution was proposed by many other conducted [12,13,14] that seek an alternative of Softmax function for classification tasks.

In this study, we aim to explore the strength of pre-trained models: AlexNet, and VGG16 that are modified by replacing their classifier with a support vector machine, which is expected to avoid overfitting that can be produced if a feedforward neural network is used. Note that AlexNet with SVM is denoted as AlexNet-SVM while VGG16 with SVM is denoted as VGG16-SVM. The networks are all trained and tested on a database for the task of iris melanocytic tumors diagnosis, which includes unaffected and tumorous irises. We aim to investigate the generalization power of these deep learning approaches when trained and tested on a small database, consists of 1500 iris images that represent 3 different categories: iris nevus unaffected, iris cysts, and iris melanocytic tumors of all its types.

It is also our aim to compare the performance of those modified models and the originals networks which use neural network classifier in the task of iris tumors classification. This comparison can prove whether the use of SVM prevents the drawbacks of using a neural network and helps in boosting the performance of the employed deep models.

### Related works

A few numbers of studies were conducted concerning the identification of such medical condition i.e., classifying the iris conditions including iris melanocytic tumors. Oyebade et al.,[15] employed CNN and deep belief network (DBN) for the identification of iris nevus, which can transform into tumor. The authors trained and tested their using different CNN, each with different learning rate. Moreover, DBN was trained and tested using different learning rates and number of hidden neurons. As a result, authors claimed that DBN outperformed the CNN in terms of accuracy, where DBN scored 93.67% while CNN reached 93.35%.

Another study by Oyebade et al.,[16] was also conducted for the same classification task but using different neural networks. This study uses backpropagation neural network (BPNN), support vector machine (SVM), and radial basis function network (RBFN) for the diagnosis of iris nevus. Experimentally, networks performed differently during training and testing using same number of images. However, the BPNN achieved the highest recognition rate as the authors stated.

## Materials and Methods

### Deep convolutional neural network (DCNN)

Deep Convolutional Neural Network is a machine learning system that consists of two-dimensional distinct convolution in image analysis to recreate the human neural system processes, which consists of a structure that resembles that of the visual nervous human system. Such type of networks was first designed by Rumelhart et al. [17] which employs the back propagation algorithm. Also, LeCun et al. [18] deployed a deep structure network while training the network parameters using the back propagation algorithm, the said network achieved a significant performance with high accuracy in determining handwritten digits.

Generally, DCNN can be described as a network that includes mathematical operations like convolution and subsampling which occurs inside its hidden layers. These processes grant the network the ability to learn various features of the networks which results in an automatic extraction of deep features and an effective presentation of the input data [19]. However, DCNN integrates a local connection structure in adding a weight sharing method in a bid to reduce the learning parameters, which invariably reduces the computational time and cost. This said network was successful when tested in the fields of computer vision [20], biological computation [21], medical images classification [22], etc.

Recent trends in in Artificial intelligence has motivated scientists to further improve the performance of CNN by making the networks deeper and much more feasible. This prompted scientists to create a CNN network which consists of 19 layers, the network was termed VGG-Net [23]. Also, a research by Szegedy et al. [24] designed and proposed a deeper network which has 22 layers, the network is called GoogleNet. GoogleNet was based on the structure of the CNN network, but an inception module was added in the structure. Moreover, another network which consists of 152 layers named ResNet (ResNet-152) was designed by He et al. [25].

### Transfer Learning

Data availability is a common issue in medicine. Data are usually limited and small which doesn’t help the intelligent systems to learn. Training the deep network with little amount of data can lead to overfitting which sometimes reduce the generalization capability of networks [21,26,37].

Transfer learning can be a good solution of that data deficiency issue [22,23,26,27,34]. In this research, two different pre-trained CNN models have been employed for the diagnosis of iris melanocytic tumors by classifying iris images into three different classes. These two convolutional neural networks are: AlexNet and VGG-16.

### AlexNet

Convolutional neural networks (CNN) originally since its conception according to Fukushima’s neocognitron model of 1982 [3], which was popular from Lecun et al. [18] also in Krizhevsky et al. [28]. The former used a CNN to come first in the ImageNet Large scale visual recognition challenge, thus making CNN’s a favourite amongst scientists for image classification tasks. One major advantage of using CNN for image classification is the capability of CNN to perform feature extraction of the data set and learn high level features from the dataset without human interference, making CNN a top-notch machine learning architecture in the field of image recognition. Convolutional neural network has received a wide acceptance in the field of machine vision in conducting task such as image classification, semantic segmentation, and object detection. Recently, CNN’s have also been deployed in the fields of classification and diagnosis using 2 dimensional images, images like chest x-rays, retinal images, dermoscopic skin images with promising results that goes beyond human classifications [4,5,29,30].

Fig. 1 shows the structure of the AlexNet where a Softmax neural network is supplanted by a multi-class SVM.

**Figure 1:**
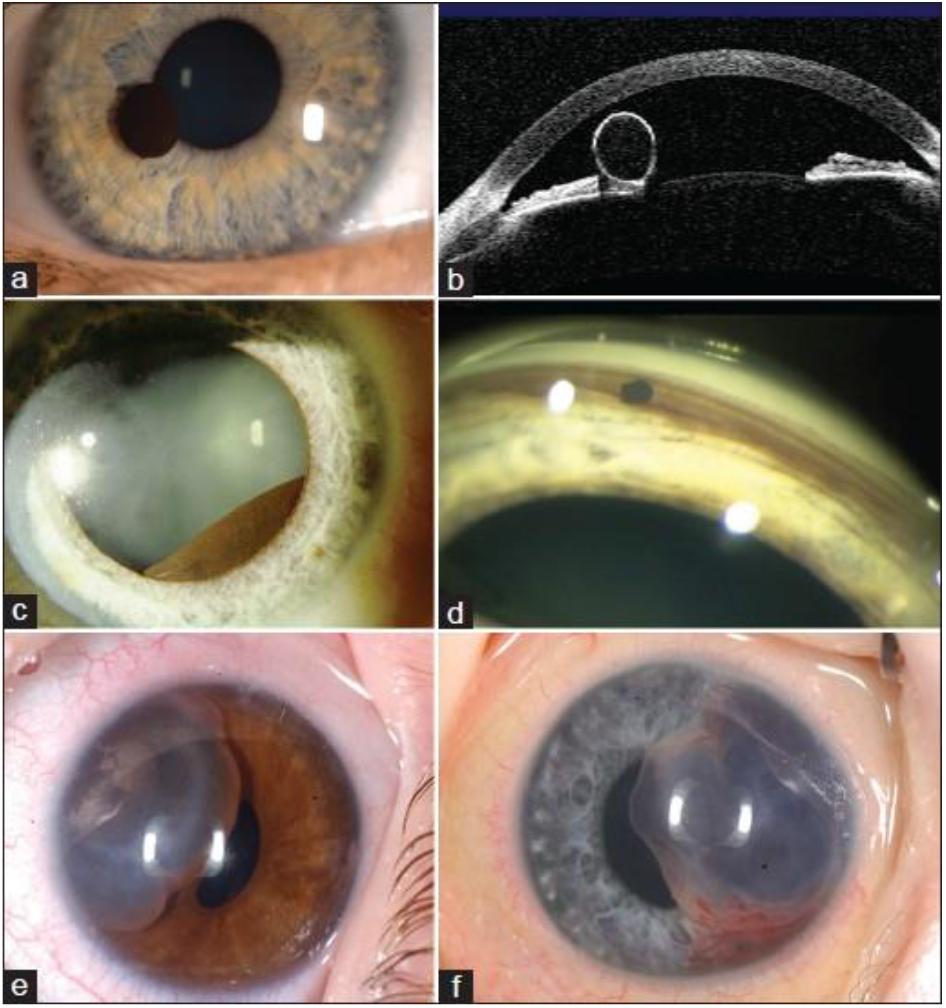
Samples of Cystic lesions. Iris cysts (a,b) Iris pigment epithelial (IPE) cyst pupillary margin seen clinically (a) and by optical coherence tomography (b), (c) IPE cyst mid-zonal, (d) IPE cyst dislodged and in the anterior chamber angle, (e) Iris stromal cyst in an infant, (f) Iris stromal cyst with hemorrhage in an infant [3]

### VGG-16

The VGG-16 is deep convolutional neural network that was proposed at ILSVRC in 2014 [23] and was able to achieve the least error rate. This network consists of 16 main layers, among them 13 have convolutional layers while the remaining have fully connected layers. Unlike the AlexNet, all layers of the convolutions that exists VGG-16 have the same filter size. Moreover, ReLU layers, max pooling layers, fully connected layers, and dropout layers are also used in the VGG-16.

**Figure 1:**
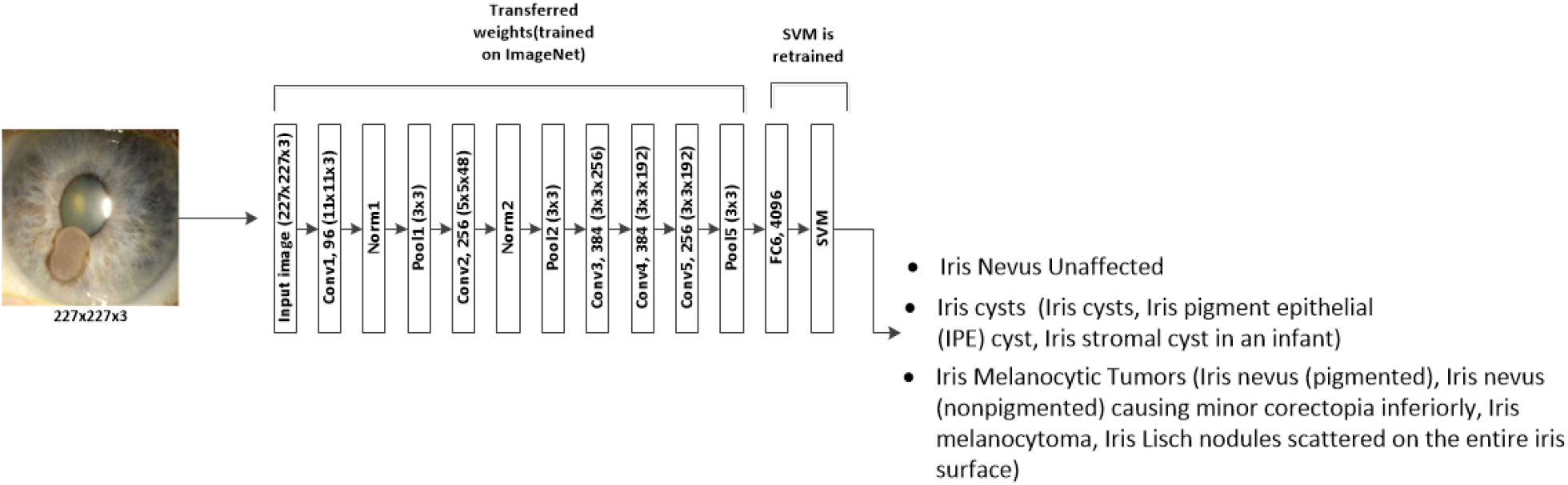
The proposed AlexNet transfer learning network for the classification of iris nevus malignancy

### Data

The four models used are trained and tested with a combination of both healthy and diseased iris images collected from two different public databases known as **The Eye Cancer Foundation, Eye Cancer Network** [35] and Miles Research [36]. The first dataset contains images of the different irises such as cysts, nevus, melanoma etc…However, the second one contains only images of the healthy or nevus unaffected iris images. Images collected from both datasets were rotated through an angular step of 15, 30, and 65 °, for the purpose of building a rotational invariance capability in the designed system and also obtaining a larger training database that would help the deep networks in learning more features of the three iris images types. The images of these databases are RGB images of different sizes; hence they were first downsampled to 227×227 and then to 224×224 pixels to fit in the input layer of the two models: AlexNet and VGGNET16, respectively. Overall, a dataset of 1500 images of the three types discussed in this work is created.

Figure 2 shows a sample of the iris images used for training and testing the re-trained models.

**Figure 2:**
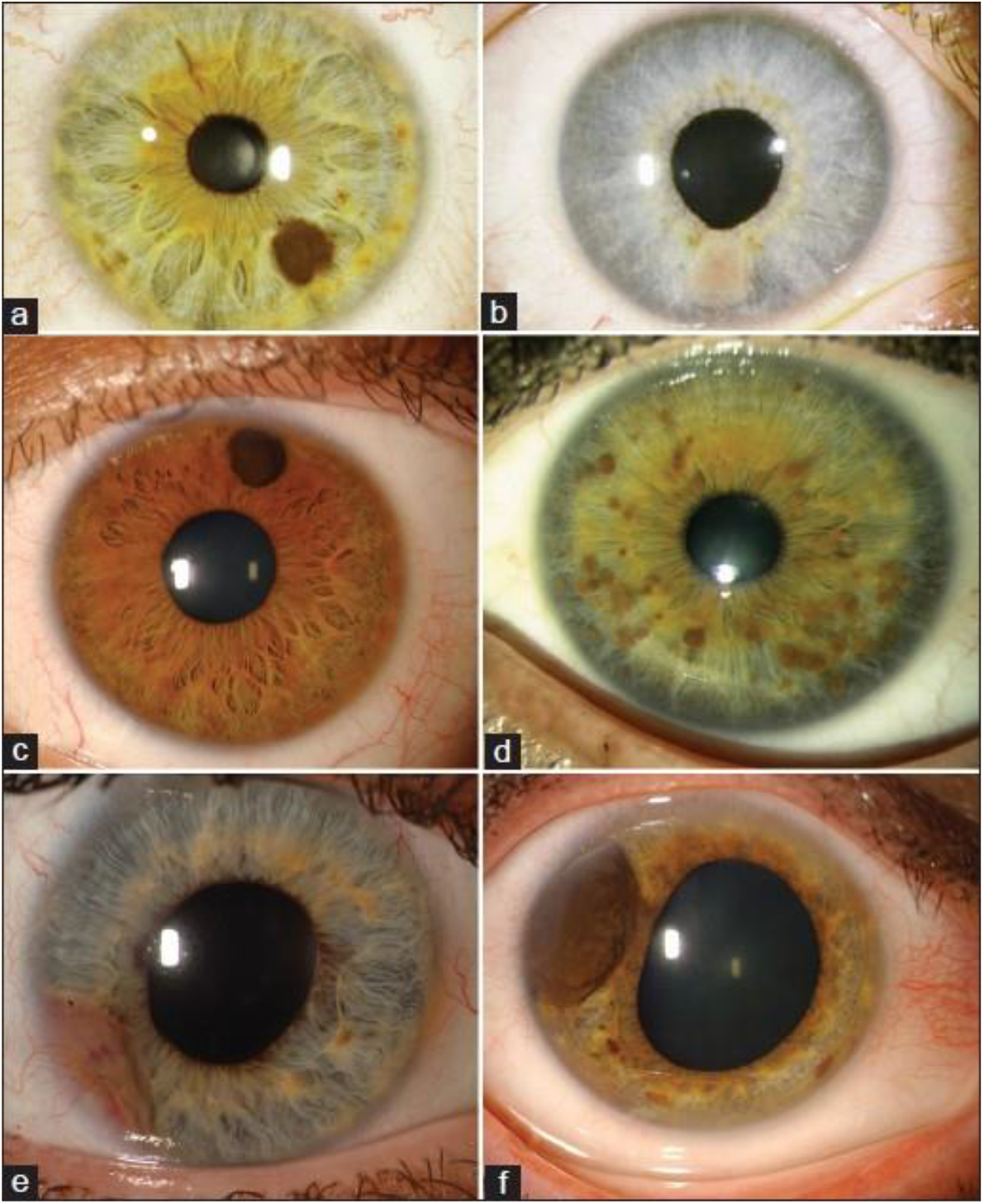
Iris melanocytic tumors (a) Iris nevus (pigmented), (b) Iris nevus (nonpigmented) causing minor corectopia inferiorly, (c) Iris melanocytoma, (d) Iris Lisch nodules scattered on the entire iris surface, (e) Iris melanoma involving the anterior chamber, (f) Iris melanoma causing corectopia [3]

**Figure 2:**
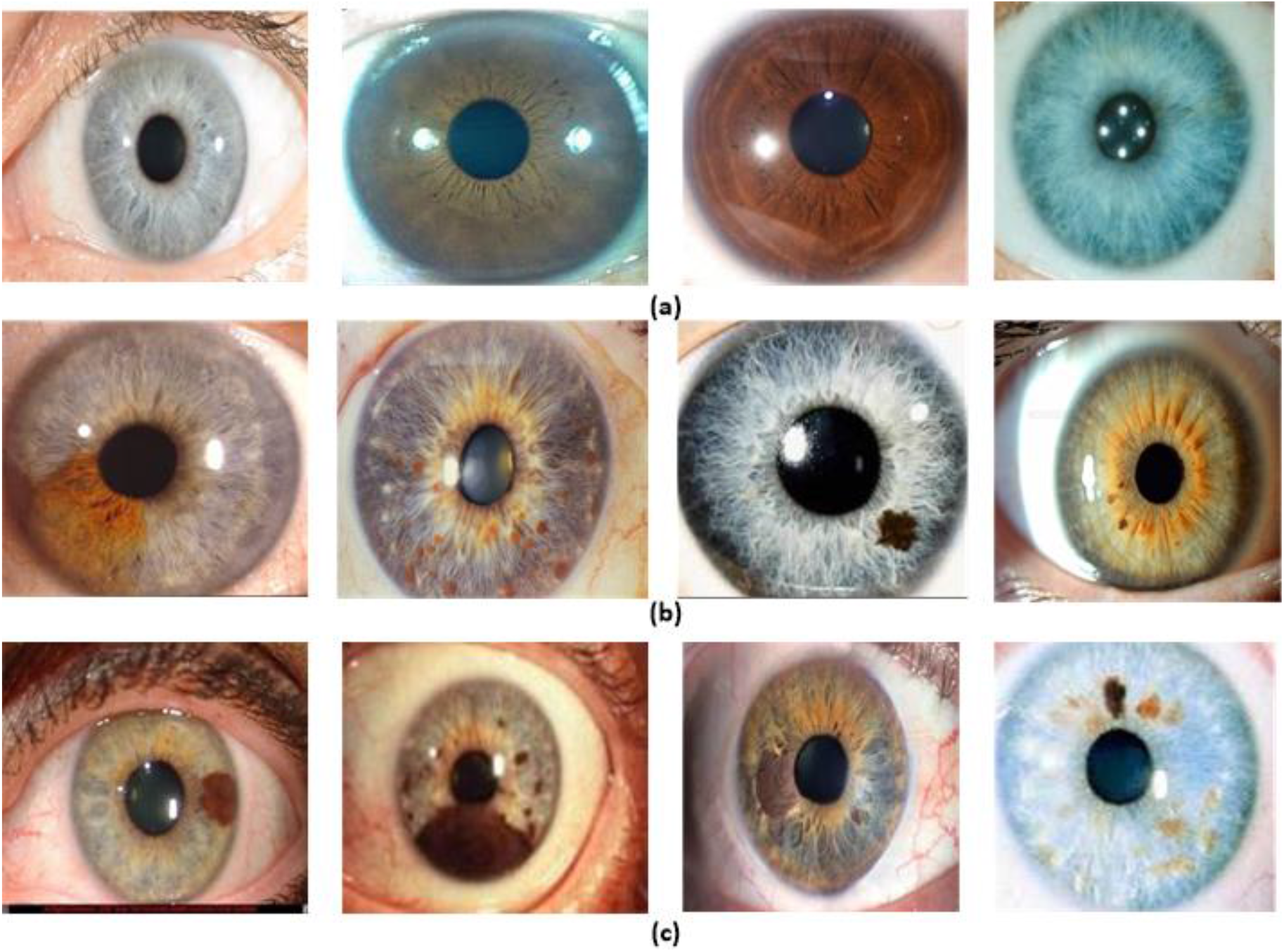
Sample of the databases training and testing images, (a) shows the nevus unaffected images, (b) shows the iris cysts images, while (c) shows the iris melanocytic images.

## Results

### Training of Pre-trained models

The pre-trained models used in this research were trained and tested on a ratio of 60% of the available data. The performance of each of the networks was then evaluated with a held-out test set of the remaining 40% of the data.

AlexNet is the first pre-trained model architecture we used, a convolutional neural network winning in the ILSVRC 2012 competition [28]. As shown in Figure 1, the network mainly consists of 5 convolutional layers denoted as CONV (CONV1 to CONV5) followed by 3 fully connected layers denoted as FC (FC6 to FC8), in addition to a Softmax activation function in the output layer (multinomial logistic regression).

Note that the openly accessible weights of the network that was trained against the ILSVRC12 was used in this transfer learning based research. As we are using a pre-trained model, its final fully connected layer FC8 was removed and a new layer was added, and it has two output neurons conforming to the three iris conditions classes. Note that the weights of this layer are modified at random. On the other hand, the other five convolutional layers are remained in the network, but their weights were frozen to act as a feature extractor. These weights are already pretrained on millions of images to extract high level features from the input data. For training, a batch size of 60 images for each iteration is used via stochastic gradient descent [31]. Also, the learning rates for the fully connected FC6, FC7, and FC8 layers were fixed at 0.001, 0.001, and 0.01, respectively during training. Consequently, this allows the network to learn much quicker to be used by the fully connected layer (FC8). Moreover, the network is fine-tuned using 60% of the available data, and 150 epochs are set to train the network.

Table 1 shows the learning parameters of the AlexNet model. It can be seen that the network has achieved a training accuracy of 94% in approximately 2 hours and 150 epochs.

**Table 1:** Models learning parameters

|  | **AlexNet** | **VGG-16** | **AlexNet-SVM** | **VGG16-SVM** |
| --- | --- | --- | --- | --- |
| Learning parameters | Values | Values | Values | Values |
| Training ratio | 60% | 60% | 60% | 60% |
| Learning rates (FC6, FC7, FC8) | 0.001, 0.001, 0.01 | 0.001, 0.001, 0.01 | - | - |
| Number of epochs | 150 | 250 | 50 | 70 |
| Training accuracy | 93.5% | 95.6% | 94.13% | 96.27% |
| Training time | 2 hours | 3.5 hours | 1.5 hours | 1 hours |
| Achieved mean square error (MSE) | 0.097 | 0.035 | 0.076 | 0.0175 |

The second employed pre-trained model in this study is the VGG16 network. As discussed earlier, this convolutional neural network is deeper than AlexNet as it consists of 13 convolutional layers denoted as CONV (CONV1 to CONV13), and 3 fully connected layers denoted as FC (FC14 to FC16), as shown in Fig.1. Note that this is also followed by a Softmax classifier as in AlexNet. Similarly, the weights of the convolutional layers of VGG-16 are fixed as they are already well trained on ImageNet to extract the higher-level abstractions of input images during ILSVRC14. Hence, only the final fully connected layers (FC14 to FC16) weights are trained, by initiating them randomly.

For training, this network is trained on 60% of the available dataset with a batch size of 65 images per iterations, which is selected to be greater than that of AlexNet, for VGG16 is deeper. Moreover, this network is trained with learning rates of 0.001, 0.001, and 0.01 for FC6, FC7, and FC8 layers, respectively. Table 1 shows the learning parameters of VGG-16. From this table, it is seen that the VGG-16 network has achieved a training accuracy of 95.9% with 250 epochs and in 3.5 hours as training time. It is also noticed that this network reached a lower mean square of 0.035 which is lower than that achieved by AlexNet. Figure 4 shows the learning curves of the VGG-16 network.

Similarly, the modified networks are trained and tested on the same images other networks used. The features extraction of these networks was handled in the same manner in the previous networks; however, the classification part here is different. An SVM is placed by taking inputs from the first fully connected layer (FC6 in AlexNet, and FC6 in VGG-16 trained to classify those features into 3 different iris categories.

As seen in table 1 these two networks are respectively trained and tested using 60% and 40% of the data. It is seen that AlexNet-SVM and VGG16-SVM required 50 and 70 epochs to reach minimum square errors of 0.076 and 0.0175, respectively. Moreover, it is noted that those networks achieved relatively higher training accuracies (94.13% and 96.27%) compared to other networks which used neural network classifier.

## Discussion

### Original models performance discussion

Upon training, both AlexNet and VGG-16 pre-trained models are tested on the same number of data, i.e. 40% of the total available data. Table 2 illustrates the performances metrics of each model during training. As shown, AlexNet achieved 93.5% accuracy, while VGG-16 was capable of achieving a higher recognition rate of 95.6%. Also, AlexNet required 150 epochs to attain such accuracy, which is lower than that needed for VGG-16 to achieve its higher accuracy. Moreover, it is noted that VGG-16 reached a lower mean square error (MSE) (0.035) than that achieved by AlexNet (0.097); however, this required longer training time. Figures 3 and 4 show the learning curves of the models. Those figures show the error variations with respect to the Epochs increasing during training of both networks, AlexNet and VGG-16, respectively. It can be seen that both networks performed well during training; however, the expansion of profundity of VGG-16 makes it increasingly hard to train, i.e. it needed longer time and more iterations to strike a good mean square error (MSE) and high performance metrics values. On the other hand, it is noteworthy that this difference in time and epochs number of VGG-16 ended up with a lower MSE than that reached by AlexNet.

**Table 2:** Original networks performance

|  | **AlexNet** | **VGG-16** |
| --- | --- | --- |
| Training accuracy | 93.5% | 95.6% |
| Testing accuracy | 89%, | 93.2% |
| MSE | 0.097 | 0.035 |

**Figure 3:**
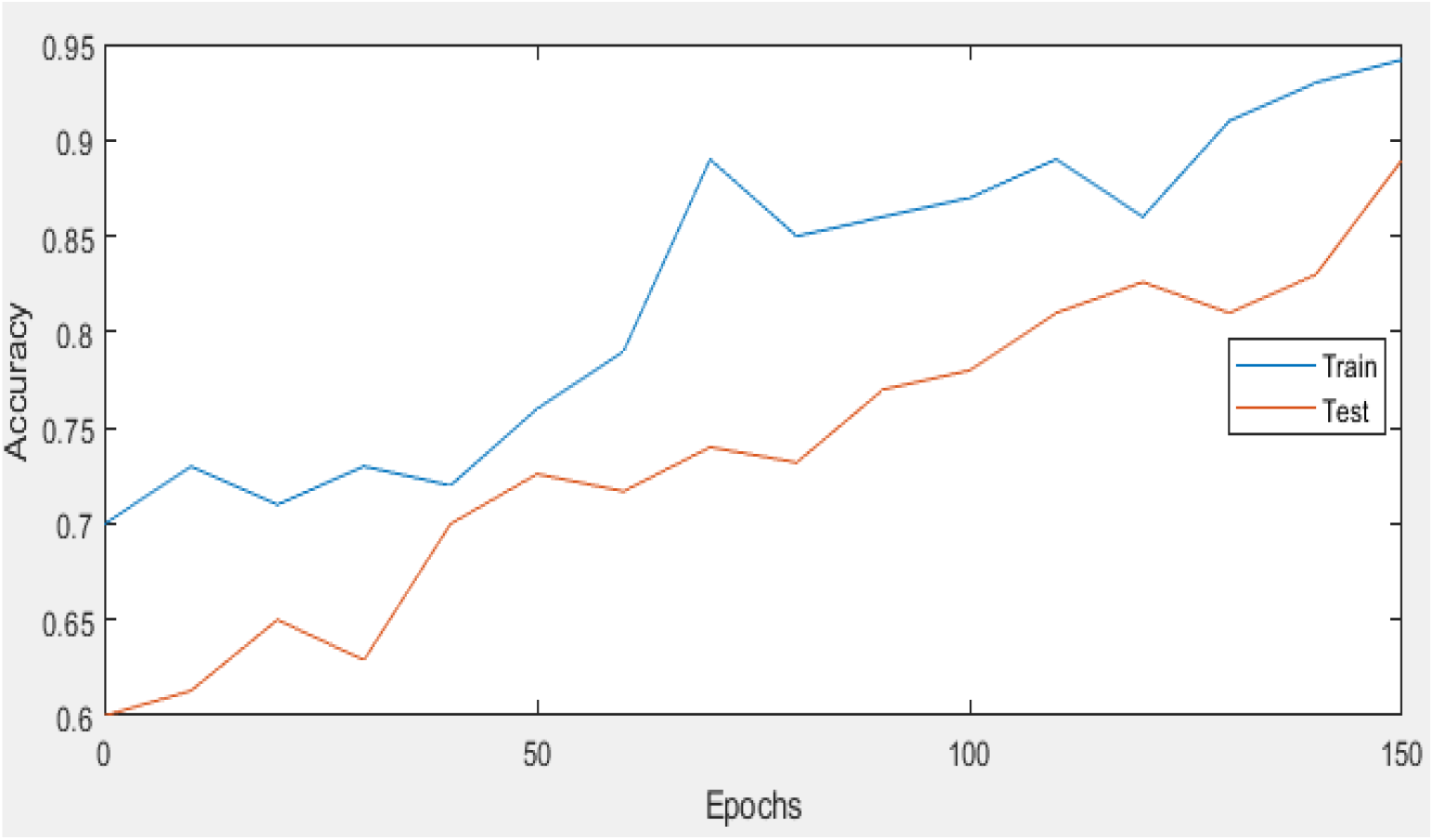
Learning curves for AlexNet

**Figure 4:**
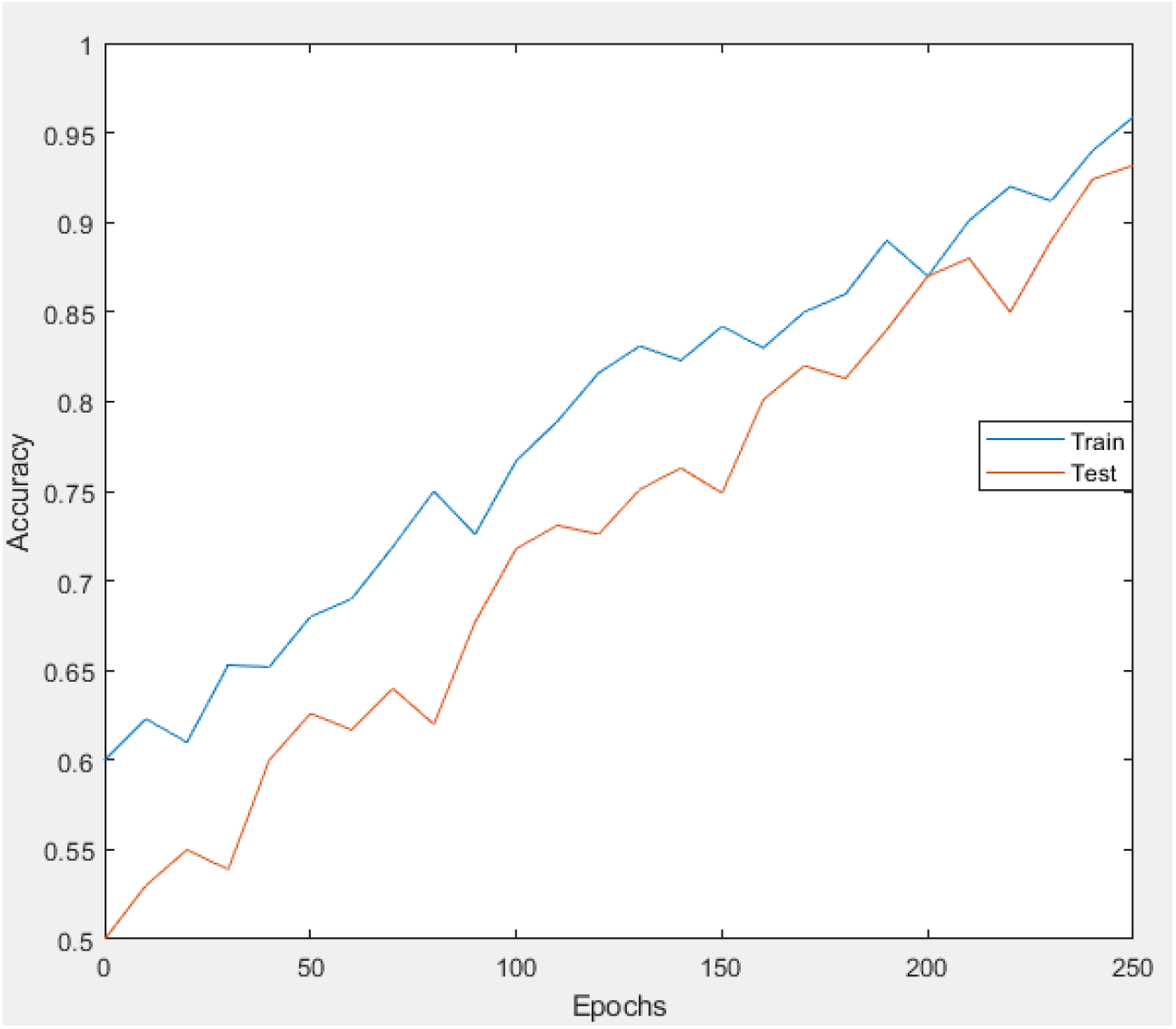
Learning curves of VGG16

For more understanding of the networks learning performance and to have comprehension into the various levels features learned by the used models, we sought to visualize the learned kernels or features in the convolutional layers.

Figure 5 shows the learned features of AlexNet, fine-tuned to classify irises into 3 different conditions. From Figure 5(a), it is seen that different levels of features are extracted at the convolutional layer 5. Figure 5(b) shows an example of one iris cyst image’s example that was investigated to visualize the activations of AlexNet in specific channels. Each square of the image on the right is an activation which is an output of a channel in the convolutional layer 1. Positive activations in this image are represented by white pixels, while black pixels depict negative or poor activations. Note that a mostly gray channel does not activate as strongly on the input image.

**Figure 5:**
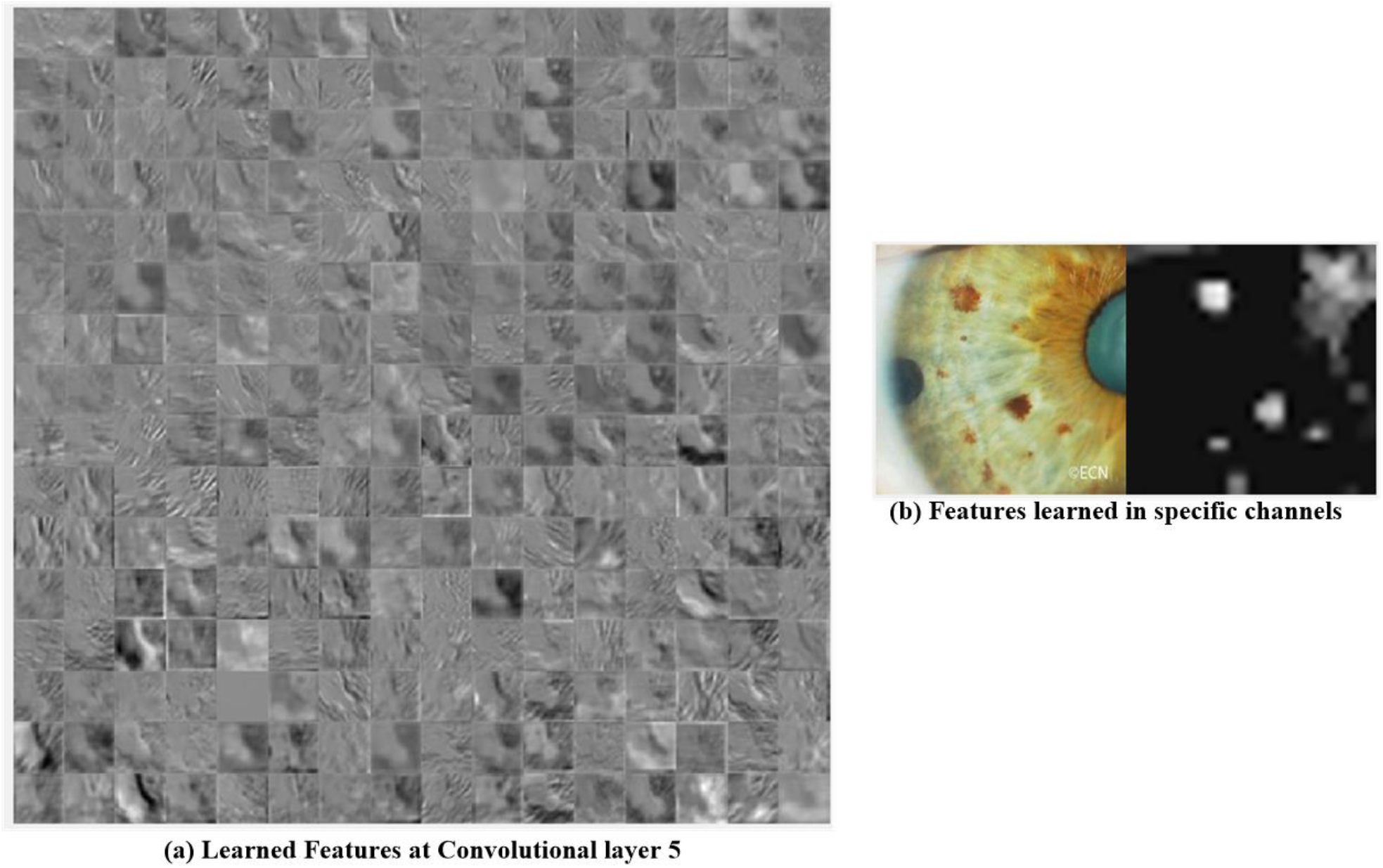
Learned features of AlexNet

Figure 6 shows the visualizations of extracted features of one iris tumor by the VGG-16 pre-trained model. It is seen that different levels of abstractions are extracted during each layer which helps the network in learning the exact and appropriate features that discriminate the 3 different classes of iris images discussed in this study.

**Figure 6:**
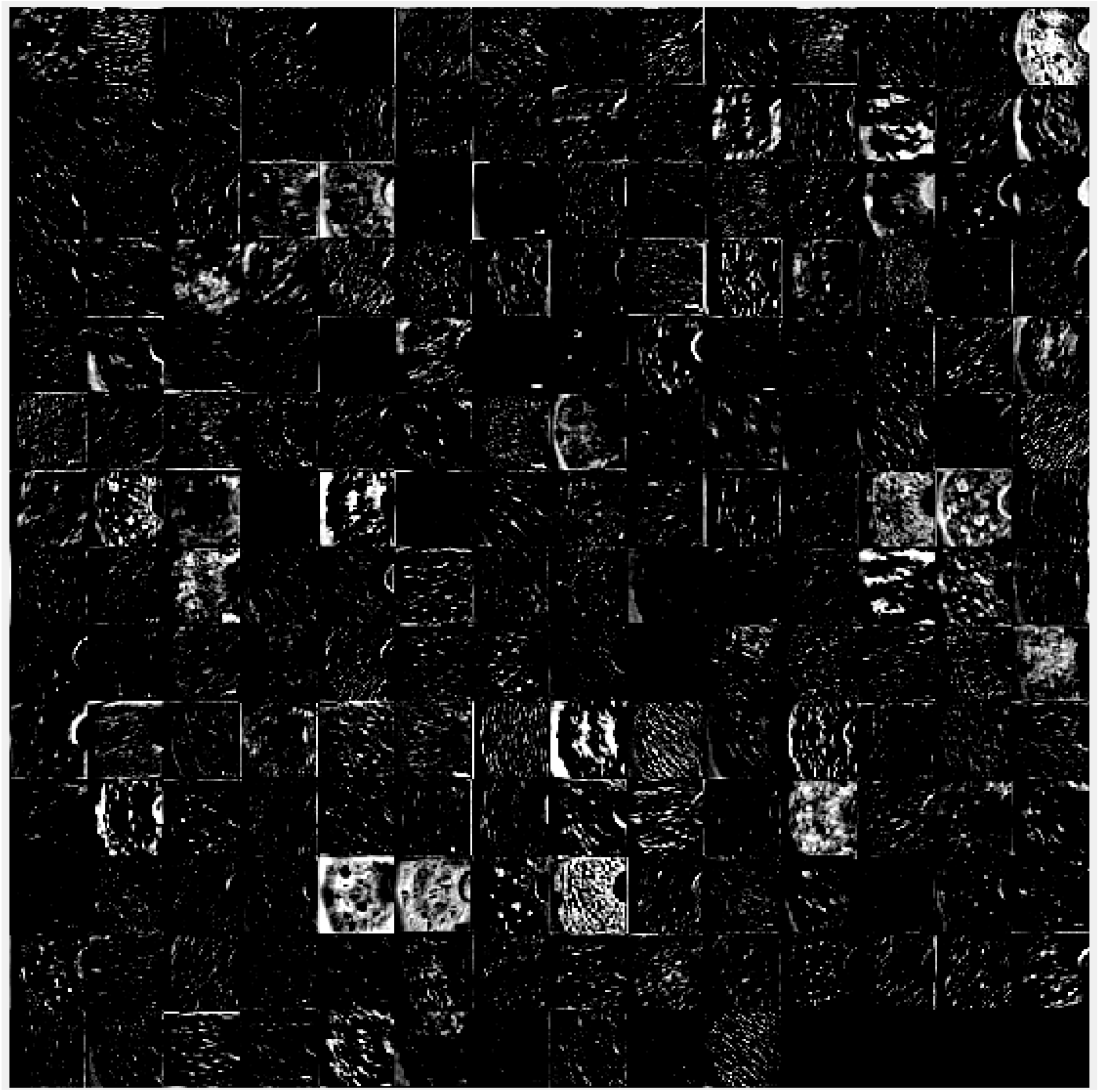
Learned features of VGG16

Table 2 shows the accuracies achieved by the employed pre-trained models for the identification of iris malignancy. Note that the networks are tested in 40% of the data. Moreover, the accuracy is calculated as in Equation 1, explained in the previous section. The table shows that VGG-16 achieved higher iris malignancy classification accuracy during testing, than that obtained by AlexNet. It is important to denote that the VGG-16 was expected to perform better than AlexNet due to the difference in depth of both networks, which allows the VGG-16 to extract more useful features than the AlexNet, and consequently this yields in a better performance. In addition, VGG-16 has achieved a lower MSE compared to that of AlexNet; but this wasn’t achieved without cost, it required a longer training time and more iterations. Furthermore, from Figure 5, it is seen that AlexNet learned some unrelated background features and information, in contrast to VGG-16. Note that this usually has negative effects on the final prediction [29,30,32].

### Modified models performance discussion

Table 3 shows the comparison of AlexNet-SVM and VGG16-SVM performances in terms of accuracy and mean square error (MSE). It is seen that VGG16-SVM outperformed the AlexNet as it achieved a higher training and testing accuracy of 91.34% and 94.13% respectively. As these two networks use the same classifier, it seems that this outperformance is due to the deeper architecture of the VGG16 network and the smaller kernel size that it uses compared to that of AlexNet. The small kernel sizes result in multiple non-linear layers and this consequently increases the depth of network, which gives it the power of extracting more complex features.

**Table 3:** Modified networks performance

|  | **AlexNet-SVM** | **VGG16-SVM** |
| --- | --- | --- |
| Training accuracy | 94.13% | 96.27% |
| Testing accuracy | 91.34% | 94.13 |
| MSE | 0.076 | 0.017 |

### Comparison between modified and original models performance

In this work, the fusion of pre-trained models (AlexNet and VGG-16) and SVM classifier was used for comparison. The combination of AlexNet and VGG-16 with SVM is more than sufficient, where AlexNet and VGG-16 perform the high-level feature extraction while SVM carried out the classification. From tables 4, it is seen that adding SVM contributed to boosting the performance of the AlexNet and VGG-16. This slight boost was not only terms of accuracy; however, SVM allows networks to converge in shorter time while reaching smaller errors.

**Table 4:** Modified and original models’ performance

|  | **AlexNet** | **VGG-16** | **AlexNet-SVM** | **VGG16-SVM** |
| --- | --- | --- | --- | --- |
| Training accuracy | 93.5% | 95.6% | 94.13% | 96.27% |
| Testing accuracy | 89%, | 93.2% | 91.34% | 94.13 |
| MSE | 0.097 | 0.035 | 0.076 | 0.017 |

Table 5 shows the accuracies achieved by all the employed networks when classifying 3 different abnormalities in the irises. Note that all experiments were performed by taking the same division of images into train and test sets of 60:40 ratio for all abnormalities.

**Table 5:** Networks performance of all the classes

|  | **Networks** |  |  |  |
| --- | --- | --- | --- | --- |
| **Classes** | **AlexNet** | **VGG-16** | **AlexNet-SVM** | **VGG16-SVM** |
| **Nevus Unaffected** | 93.56% | 94.55% | 95.18% | 97.65% |
| **Iris Cysts** | 87.95% | 89.65% | 92.68% | 90.47% |
| **Iris Melanocytic Tumors** | 74.25% | 72.23% | 88.32% | 89.63% |

**Table 6:** Performance evaluation metrics

| Network Model | AlexNet | VGG-16 | AlexNet-SVM | VGG16-SVM |
| --- | --- | --- | --- | --- |
| Accuracy | 89% | 93.2% | 91.34% | 94.13% |
| Sensitivity | 91% | 95% | 94% | 95% |
| Specificity | 84% | 89% | 87% | 90% |

### Performance Evaluation Metrics

Confusion matrix is one the most common and effective metrics to evaluate the performance of deep networks, in classification tasks [13]. Metrics that can be derived from the confusion matrix are accuracy, sensitivity, and specificity. Accuracy is the number of correctly predicted classes over all other predictions. It is a good measure to show that whether or not data are closely balanced. Sensitivity indicates the proportion of correctly classified positive data points with respect to all other positive data points; Specificity shows the proportion of correctly classified negative data points with respect to all other negative data points.

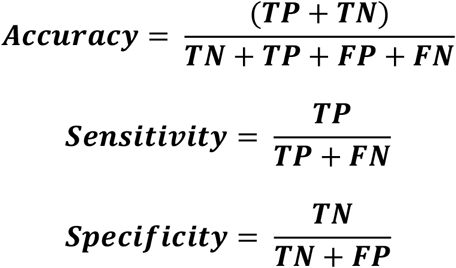

### Comparison

A comparison of the developed networks employed in this work with some earlier works is shown in Table 5. Firstly, it is seen that the employed pre-trained CNNs with SVM achieved higher accuracies during testing, compared to various deep networks tested, which might be owed to their powerful efficiency in extracting the important features from input images. The convolutional neural networks (CNNs) employed within this work achieved higher accuracies than other earlier work that used a BPNN features, which achieved highest comparison compared to SVM and RBFN [16]. Furthermore, it is important to note that our employed networks gained a better generalization capability compared to those other networks employed for iris melanocytic tumor classification such as DBN [15].

| Parameters | AlexNet | VGG-16 | AlexNet-SVM | VGG16-SVM | CNN<br>[15] | DBN<br>[15] |
| --- | --- | --- | --- | --- | --- | --- |
| Testing Accuracy | 89%, | 93.2% | 91.34% | 94.13 | 93.3% | 93.67% |

## Conclusion

In this research, transfer learning which is based on neural networks were employed. AlexNet and VGG-16 are both used. Their features learned on a source task are transferred to a new task, eyes dataset, in order to determine the classification of iris cases into three different classes: Unaffected nevus, iris cysts, and iris melanocytic tumors which include different types of eye tumors. Results showed that VGG-16, a well-designed and deeper architecture of adequate complexity, was capable of achieving considerably higher classification accuracy when distinguishing between three different classes, as compared to that of AlexNet. Furthermore, VGG-16 network learned features visualization demonstrates that mid and high-level features are learned effectively by the model.

It can be concluded that transfer of knowledge learned by a well-trained convolutional network to learn a new classification task is possible, regardless of small margins of errors when trained using a relatively small database. Moreover, it is important to conclude that depth of the convolutional networks can help in a better understanding and analysis of the images, which helps in extraction different levels of abstraction through the network’s convolutions and pooling layers. Furthermore, it is found that replacing Softmax neural network with SVM contributes to a slight boosting of the network performance and reducing its training time. Lastly, our results can show that applying deep pre-trained CNN models combined with SVM instead of Softmax, for the problem of iris melanocytic tumors analysis, its favorable in a way that related or confusing iris images can be recognized or correctly classified with good recognition rates. Convolutional neural networks benefit from the hierarchical learning and representations of data which have been found to contribute to obtain superior performances in various applications and therefore in this research.

## Data Availability

Data is available upon request.

## References

[1] Shields, C. L., Kancherla, S., Patel, J., Vijayvargiya, P., Suriano, M. M., Kolbus, E., … & Zhang, Z. (2012). Clinical survey of 3680 iris tumors based on patient age at presentation. Ophthalmology, 119(2), 407–414.

[2] Shields JA, Shields CL. Intraocular Tumors. An Atlas and Textbook. 2nd ed. Philadelphia: Lippincott Williams and Wilkins; 2008. p. 3–58.

[3] Shields, C. L., Shields, P. W., Manalac, J., Jumroendararasame, C., & Shields, J. A. (2013). Review of cystic and solid tumors of the iris. Oman journal of ophthalmology, 6(3), 159.

[4] Oyedotun, O., & Khashman, A. (2017). Iris nevus diagnosis: convolutional neural network and deep belief network. Turkish Journal of Electrical Engineering & Computer Sciences, 25(2), 1106–1115.

[5] Abiyev, R. H., & Ma’aitah, M. K. S. (2018). Deep convolutional neural networks for chest diseases detection. Journal of healthcare engineering, 2018.

[6] Albarqouni, S., Baur, C., Achilles, F., Belagiannis, V., Demirci, S., & Navab, N. (2016). Aggnet: deep learning from crowds for mitosis detection in breast cancer histology images. IEEE transactions on medical imaging, 35(5), 1313–1321.

[7] Bengio, Y., Lamblin, P., Popovici, D., & Larochelle, H. (2007). Greedy layer-wise training of deep networks. In Advances in neural information processing systems (pp. 153–160).

[8] Kaymak, S., Helwan, A., & Uzun, D. (2017). Breast cancer image classification using artificial neural networks. Procedia computer science, 120, 126–131.

[9] Russakovsky, O., Deng, J., Su, H., et al. “ImageNet Large Scale Visual Recognition Challenge.” International Journal of Computer Vision (IJCV). Vol 115, Issue 3, 2015, pp. 211–252.

[10] Bishop, C.M. (2006) Pattern Recognition and Machine Learning, Springer-Verlag, New York, USA.

[11] Oyedotun, O. K., Olaniyi, E. O., & Khashman, A. (2017). A simple and practical review of over-fitting in neural network learning. International Journal of Applied Pattern Recognition, 4(4), 307–328.

[12] Abien Fred Agarap. 2017. A Neural Network Architecture Combining Gated Recurrent Unit (GRU) and Support Vector Machine (SVM) for Intrusion Detection in Network Traffic Data. arXiv preprint 1709.03082 (2017).

[13] Abdulrahman Alalshekmubarak and Leslie S Smith. 2013. A novel approach combining recurrent neural network and support vector machines for time series classification. In Innovations in Information Technology (IIT), 2013 9th International Conference on. IEEE, 42–47.

[14] Yichuan Tang. 2013. Deep learning using linear support vector machines. arXiv preprint 1306.0239 (2013).

[15] Oyedotun, O., & Khashman, A. (2017). Iris nevus diagnosis: convolutional neural network and deep belief network. Turkish Journal of Electrical Engineering & Computer Sciences, 25(2), 1106–1115.

[16] Oyedotun, O. K., Olaniyi, E. O., Helwan, A., & Khashman, A. (2014). Decision support models for iris nevus diagnosis considering potential malignancy. International Journal of Scientific & Engineering Research, 5(12), 421.

[17] D.E. Rumelhart, G.E. Hinton, R.J. Williams, Learning Internal Representations By Error Propagation, MIT Press, 1988.

[18] Y. LeCun, L. Bottou, Y. Bengio, P. Haffner, Gradient-based learning applied to document recognition, Proc. IEEE 86 (1998) 2278–2324.

[19] Y. LeCun, Y. Bengio, G. Hinton, Deep learning, Nature 521 (2015) 436–444.

[20] Oyedotun, O. K., & Khashman, A. (2017). Deep learning in vision-based static hand gesture recognition. Neural Computing and Applications, 28(12), 3941–3951.

[21] Rios, A., & Kavuluru, R. (2015, September). Convolutional neural networks for biomedical text classification: application in indexing biomedical articles.In Proceedings of the 6th ACM Conference on Bioinformatics, Computational Biology and Health Informatics (pp. 258–267). ACM.

[22] Wang, X., Peng, Y., Lu, L., Lu, Z., Bagheri, M., & Summers, R. M. (2017). ChestX-ray8: Hospital-scale Chest X-ray Database and Benchmarks on Weakly-Supervised Classification and Localization of Common Thorax Diseases. arXiv preprint 1705.02315.

[23] K. Simonyan, A. Zisserman, Very Deep Convolutional Networks for Large-Scale Image Recognition, 1409.1556, 2014.

[24] Szegedy, C., Liu, W., Jia, Y., Sermanet, P., Reed, S., Anguelov, D., … & Rabinovich, A. (2015, June). Going deeper with convolutions. Cvpr.

[25] K. He, X. Zhang, S. Ren, J. Sun, Deep residual learning for image recognition, in: 2016 IEEE Conference on Computer Vision and Pattern Recognition (CVPR), 2016, pp. 770–778.

[26] M. Long, Y. Cao, J. Wang, M.I. Jordan, Learning transferable features with deep adaptation networks, Comput. Sci. (2015) 97–105.

[27] Cheng, P. M., & Malhi, H. S. (2017). Transfer learning with convolutional neural networks for classification of abdominal ultrasound images. Journal of digital imaging, 30(2), 234–243.

[28] Krizhevsky, A., Sutskever, I., & Hinton, G. E. (2012). Imagenet classification with deep convolutional neural networks. In Advances in neural information processing systems (pp. 1097–1105).

[29] Helwan, A., El-Fakhri, G., Sasani, H., & Uzun Ozsahin, D. (2018). Deep networks in identifying CT brain hemorrhage. Journal of Intelligent & Fuzzy Systems, (Preprint), 1–1.

[30] N. Srivastava, G. Hinton, A. Krizhevsky, I. Sutskever, and R. Salakhutdinov, “Stacked denoising autoencoder and dropout together to prevent overfitting in deep neural network,” Journal of Machine Learning Research, vol. 15, pp. 1929–1058, June 2014.

[31] R.G.J. Wijnhoven, P.H.N. de With, Fast training of object detection using stochastic gradient descent, in: International Conference on Pattern Recognition (ICPR), 2010, pp. 424–427.

[32] Yu, W., Yang, K., Bai, Y., Xiao, T., Yao, H., & Rui, Y. (2016). Visualizing and comparing AlexNet and VGG using deconvolutional layers. In Proceedings of the 33 rd International Conference on Machine Learning.

[33] Bar, Y., Diamant, I., Wolf, L., Lieberman, S., Konen, E., & Greenspan, H. (2015, April). Chest pathology detection using deep learning with non-medical training. In Biomedical Imaging (ISBI), 2015 IEEE 12th International Symposium on (pp. 294–297).

[34] Islam, M. T., Aowal, M. A., Minhaz, A. T., & Ashraf, K. (2017). Abnormality Detection and Localization in Chest X-Rays using Deep Convolutional Neural Networks. arXiv preprint 1705.09850.

[35] The Eye Cancer Foundation, Eye Cancer Network (http://www.eyecancer.com/research/image-gallery/12/iris-tumors)

[36] Miles Research http://milesresearch.com/main/links.html

